# Factors informing healthcare workers’ willingness to work during the COVID-19 pandemic

**DOI:** 10.1101/2021.03.21.21254048

**Authors:** Marta Malesza

## Abstract

**Objective:** The COVID-19 pandemic represents a substantial challenge to healthcare workers. Exploring the determinants of their willingness to work is crucial to ensuring hospital function during the outbreak. Hence, this study examines the factors affecting the motivation and hesitation of health workers in the face of the COVID-19 pandemic in Poland.

**Study Design:** An online, anonymous survey was carried out among Polish healthcare workers during the COVID-19 pandemic.

**Method:** The respondents were asked about their demographic characteristics, stress-related factors, and their self-reported motivation and hesitation to work. The responses were gathered during September-December 2020.

**Results:** 912 valid responses were obtained. Of these, 22.8% (N = 208) respondents reported being strongly motivated to work while 37.8% (N = 345) expressed strong hesitation. The participants’ demographic characteristics and their responses to the stress-related questions were assigned to four categories depending on the odds ratios of motivation and hesitation. While some factors were linked to enhanced motivation and reduced hesitation, others solely affected either motivation or hesitation, and yet others had a positive impact on both.

**Conclusion:** Overall, the study indicates that in order to increase hospital workers’ motivation and decrease their hesitation, they must be made to feel protected by both their hospitals and local and national authorities.

## Background

Health workers’ professionalism is frequently been challenged, no more so than in the context of epidemics, e.g. Ebola [1], SARS [2], and it is no different during the COVID-19 pandemic [3]. In order to ensure that hospitals can continue to function even under such difficult circumstances, it is crucial to determine which factors inform health workers’ professional attitudes in such events. In particular, the public emergency caused by the SARS epidemic highlighted the importance of such an investigation, leading to several studies that questioned health workers on their potential response to hypothetical pandemics. In German hospitals, 28% of health workers reported a hesitation to work in a pandemic stemming from their concern for themselves and their families [4]. Among US public health workers presented with a hypothetical influenza pandemic, 46.2% stated they would be unwilling to work [5], while 21.7% said the same of a SARS pandemic [6]. Meanwhile, 27.7% of primary care physicians in Singapore questioned about their attitudes towards an avian influenza pandemic stated they would not attend to infected individuals [7]. Family physicians in Canada similarly responded, stating they would not work in a pandemic even when asked to do so by the public health authorities [8]. The research has shown that, in general and irrespective of nationality, up to one-third of health workers are likely to display hesitation when asked to work during a pandemic.

Healthcare workers are usually expected to do their duty in caring for the sick, despite this putting them at great personal risk – a duty that forms the foundations of the professional health workers’ codes of conduct [9]. However, this duty is not unconditional as health workers are likely to have numerous other duties, e.g., to their health or that of their families [10]. This duty of healthcare workers is an emerging focus point in light of the COVID-19 pandemic – an international crisis that is having a devasting and long-lasting impact, including in Poland, where health services are becoming increasingly overburdened. The situation is unlikely to improve during the cold season, which brings with it many further complications, not least influenza.

Health workers serve on a dual frontier – they must bear the burden of caring for the sick while also exposing themselves to potential infection. However, their contribution is crucial to the effectiveness of the public health response to the pandemic as maintaining proper standards of healthcare – whether for COVID-19 patients or conventional patients – requires them to remain healthy and come to work, irrespective of the risks involved. The existing research indicates that an influenza pandemic may see substantial absenteeism in accordance with its severity and the extent of its adverse effects [11]. Hence, planning for pandemics tends to rely on tackling hurdles to health workers’ ability to work, e.g. granting flexible working hours or transport in the event they are assigned to a different hospital. Such planning makes the assumption that overcoming these hurdles to the ability to work will increase the willingness to work, i.e. that willingness and ability to work are separate yet complementary. Yet, developing practical solutions may not inevitably lead to an increase in willingness, as this is likely to originate from several determinants, e.g. health workers’ perception of being taken into consideration in preparedness planning in addition to a plethora of social or family issues; thus, health workers’ willingness to risk themselves at work during a crisis are likely to be more influenced by these [9, 11-13].

Research in hospitals has indicated that workers’ perceptions of their efficacy and the threat to themselves substantially shape their willingness to work in a crisis. Meanwhile, there are both incentives and disincentives that inform health workers’ professional conduct. Based on the above, the conflict between incentives (e.g. motivation) and disincentives (e.g. hesitation) is likely to affect health workers’ willingness to work. Therefore, gaining an insight into what affects motivation and hesitation in health workers is crucial to maintaining their willingness and thereby ensuring that hospitals can function smoothly during emergencies. Against this backdrop, this study examines the determinants of motivation and hesitation to work during the COVID-19 pandemic among health employees working in Polish hospitals.

## Methods

The study population comprised healthcare workers from three Polish hospitals that began treating COVID-19 patients in March 2020, which was when the first local case of infection was recorded in the country. Anonymously self-administered online questionnaire surveys were emailed to workers on 12^th^ September 2020. The participants were asked to send their answers online until 31^st^ December 2020. The case numbers grew steadily by around 20,000 to 25,000 each day in Poland during the survey period [14]. In total 1315 healthcare workers received questionnaires by internal email or mail. 912 individuals provided their answers (response rate = 69.3%). The employments of hospital specialists were grouped into three categories: (1) clinical staff (doctors and nurses; n = 402); (2) clinical technical/support staff (e.g., radiological technologists, laboratory technicians; n = 388); and (3) non-clinical staff (office workers, administrators, clinical clerks, guards; n = 122).

### Survey content

The survey measured the respondents’ stress-related factors, on the one hand, and their self-reported motivation and hesitation to work, on the other hand. Items taken from prior studies conducted in the context of the H1N1 and SARS outbreaks [4, 6, 7, 15, 16, 17] were used to measure the former. Specifically, the respondents were asked about their stress due to anxiety about contracting the disease, contracting it during their commute, and passing the disease to family members. They were also asked regarding their lack of knowledge of the disease’s virulence and infectiousness and about methods of protection and prevention; regarding their feelings of being protected by local and national governments and by the hospitals (e.g., reasonable action to prevent infection, including supporting health workers in not becoming patients, caring for those who contracted the disease, adequately compensating families of those who died due to being infected at work, and mitigating malpractice suits in high-risk emergency settings); regarding their feelings of isolation and avoidance by others and of being forced to work; and regarding the burden of extra work, of a shift in the type of work, and of the need to find child care (e.g., no available nursery). Finally, the respondents were asked to report their mental and physical exhaustion and experiences of insomnia and elevated mood. The respondents rated their answers to these items on a 4-point Likert scale (0 = “never”; 1 = “rarely”; 2 = “sometimes”; 3 = “always”). A similar 4-point Likert scale was used to measure the respondents’ responses concerning their feelings of motivation and hesitation to work (items: ‘’ I feel motivated/hesitated to work’’).

### Data analysis

The responses to both parts of the questionnaire were dichotomized into weak (i.e., having a score of no more than two) and strong (i.e., all others) responses. To examine the link between the demographic statistics, the stress factors, and motivation and hesitation to work, the odds ratios (OR) were calculated using bivariate and multivariate logistic regression models, adjusted for age, sex, job, and place of work.

## Results

A total of 912 participants provided their answers. 208 (22.8%) of these 912 respondents reported having the motivation to work, while 345 (37.8) expressed hesitation. As presented in Table 1, the results for the demographical statistics and ORs reveal that female respondents reported less motivation and more hesitation than male respondents. Meanwhile, respondents in the 31-40 age group were more motivated than those in the 21-30 group, although there was no significant difference in their reported hesitation. Respondents in the 41-50 and 51-60 age groups expressed lower hesitation and higher motivation, while individuals in the 61-65 age group showed greater hesitation and reduced motivation to work. Working in technical and administrative support was linked to having more motivation than clinical staff, with no significant difference regarding hesitation. Respondents in high-risk settings showed more motivation than those from low-risk settings, again with no significant difference in hesitation.

**Table 1.**
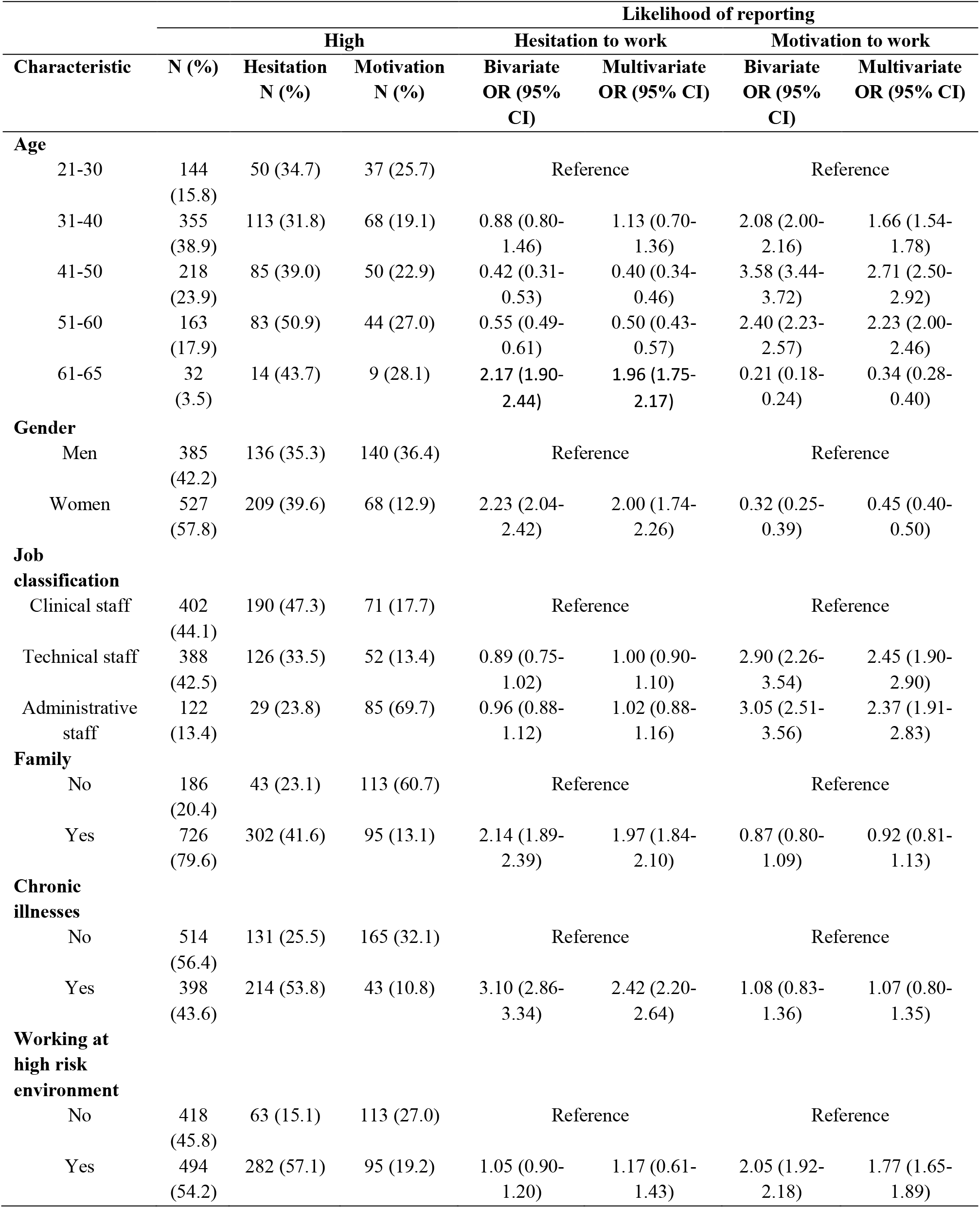
Demographic characteristics of the study population and likelihood of reporting hesitation and motivation to work

| Characteristic | Likelihood of reporting |  |  |  |  |  |  |
| --- | --- | --- | --- | --- | --- | --- | --- |
|  | N (%) | High |  | Hesitation to work |  | Motivation to work |  |
|  |  | Hesitation<br>N (%) | Motivation<br>N (%) | Bivariate<br>OR (95%<br>CI) | Multivariate<br>OR (95% CI) | Bivariate<br>OR (95%<br>CI) | Multivariate<br>OR (95% CI) |
| Age |  |  |  |  |  |  |  |
| 21-30 | 144<br>(15.8) | 50 (34.7) | 37 (25.7) | Reference |  | Reference |  |
| 31-40 | 355<br>(38.9) | 113 (31.8) | 68 (19.1) | 0.88 (0.80-1.46) | 1.13 (0.70-1.36) | 2.08 (2.00-2.16) | 1.66 (1.54-1.78) |
| 41-50 | 218<br>(23.9) | 85 (39.0) | 50 (22.9) | 0.42 (0.31-0.53) | 0.40 (0.34-0.46) | 3.58 (3.44-3.72) | 2.71 (2.50-2.92) |
| 51-60 | 163<br>(17.9) | 83 (50.9) | 44 (27.0) | 0.55 (0.49-0.61) | 0.50 (0.43-0.57) | 2.40 (2.23-2.57) | 2.23 (2.00-2.46) |
| 61-65 | 32<br>(3.5) | 14 (43.7) | 9 (28.1) | 2.17 (1.90-2.44) | 1.96 (1.75-2.17) | 0.21 (0.18-0.24) | 0.34 (0.28-0.40) |
| Gender |  |  |  |  |  |  |  |
| Men | 385<br>(42.2) | 136 (35.3) | 140 (36.4) | Reference |  | Reference |  |
| Women | 527<br>(57.8) | 209 (39.6) | 68 (12.9) | 2.23 (2.04-2.42) | 2.00 (1.74-2.26) | 0.32 (0.25-0.39) | 0.45 (0.40-0.50) |
| Job<br>classification |  |  |  |  |  |  |  |
| Clinical staff | 402<br>(44.1) | 190 (47.3) | 71 (17.7) | Reference |  | Reference |  |
| Technical staff | 388<br>(42.5) | 126 (33.5) | 52 (13.4) | 0.89 (0.75-1.02) | 1.00 (0.90-1.10) | 2.90 (2.26-3.54) | 2.45 (1.90-2.90) |
| Administrative<br>staff | 122<br>(13.4) | 29 (23.8) | 85 (69.7) | 0.96 (0.88-1.12) | 1.02 (0.88-1.16) | 3.05 (2.51-3.56) | 2.37 (1.91-2.83) |
| Family |  |  |  |  |  |  |  |
| No | 186<br>(20.4) | 43 (23.1) | 113 (60.7) | Reference |  | Reference |  |
| Yes | 726<br>(79.6) | 302 (41.6) | 95 (13.1) | 2.14 (1.89-2.39) | 1.97 (1.84-2.10) | 0.87 (0.80-1.09) | 0.92 (0.81-1.13) |
| Chronic<br>illnesses |  |  |  |  |  |  |  |
| No | 514<br>(56.4) | 131 (25.5) | 165 (32.1) | Reference |  | Reference |  |
| Yes | 398<br>(43.6) | 214 (53.8) | 43 (10.8) | 3.10 (2.86-3.34) | 2.42 (2.20-2.64) | 1.08 (0.83-1.36) | 1.07 (0.80-1.35) |
| Working at<br>high risk<br>environment |  |  |  |  |  |  |  |
| No | 418<br>(45.8) | 63 (15.1) | 113 (27.0) | Reference |  | Reference |  |
| Yes | 494<br>(54.2) | 282 (57.1) | 95 (19.2) | 1.05 (0.90-1.20) | 1.17 (0.61-1.43) | 2.05 (1.92-2.18) | 1.77 (1.65-1.89) |

Contrary, people with chronic illnesses and individuals with family showed greater hesitation than those without chronic illnesses and living without family, with no significant difference in motivation.

Table 2 presents the relationships between the stress factors and healthcare workers’ willingness to work. Factors “being protected by government” and “being protected by hospital’’ were significantly linked to higher motivation and lower hesitation. The item “elevated mood” revealed more motivation without a corresponding significant difference in hesitation. Next, the items ‘‘burden of change of quality of work’’, ‘‘burden of increased quantity of work’’, “burden of child care including lack of nursery”, ‘‘lack of knowledge about infection’’, ‘‘lack of knowledge about prevention and protection’’, ‘‘feeling of being avoided by others’’, ‘‘feeling of being isolated’’ and ‘‘feeling of being stigmatized’’ showed greater hesitation without a corresponding significant differences in motivation. Meanwhile, having both higher motivation and higher hesitation were linked to ‘‘feeling of having no choice but to work due to obligation’’, “anxiety about being infected”, “anxiety of being infected during commuting”, “anxiety about infecting family”, “physical exhaustion” and “mental exhaustion”.

**Table 2.**
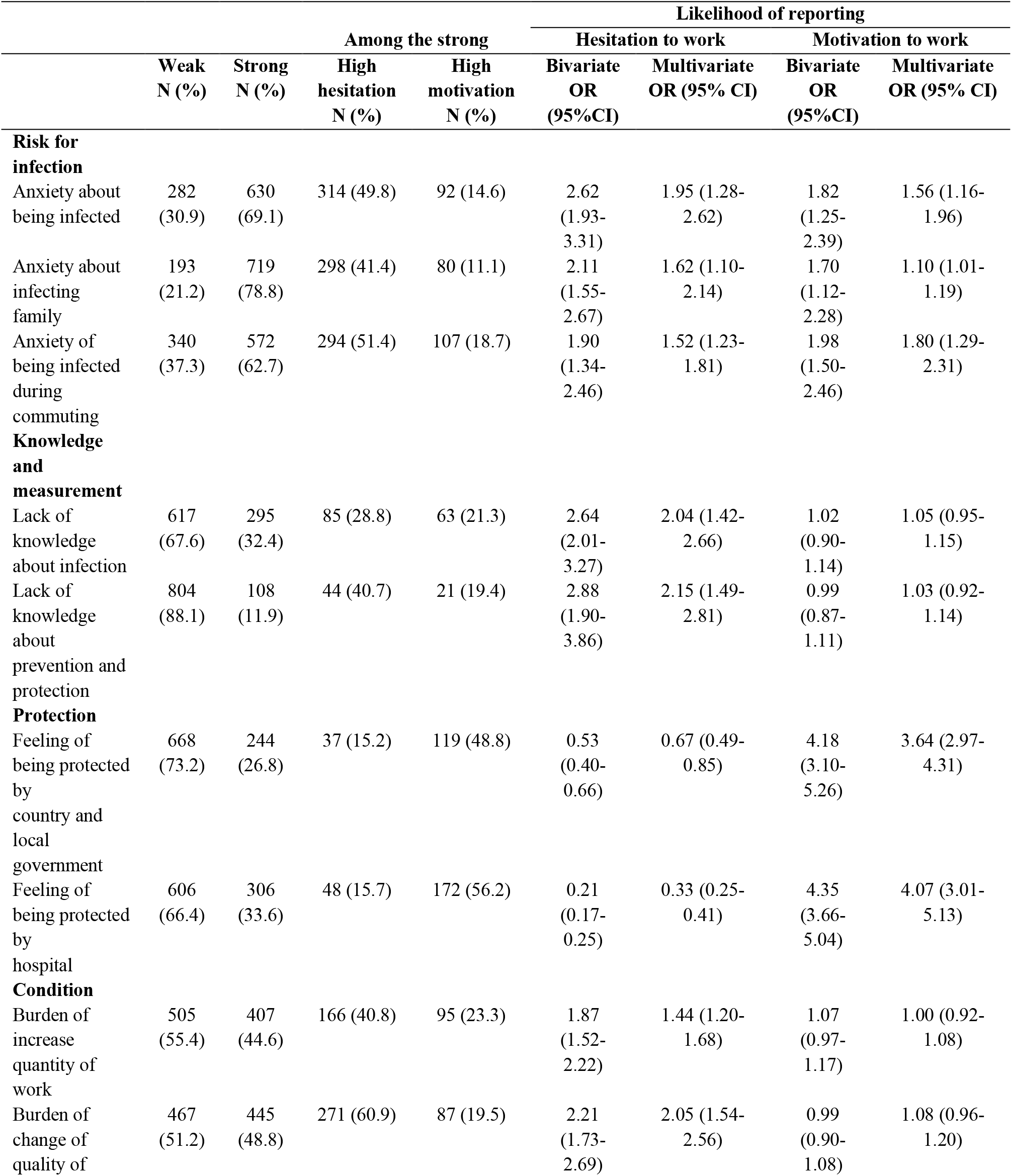
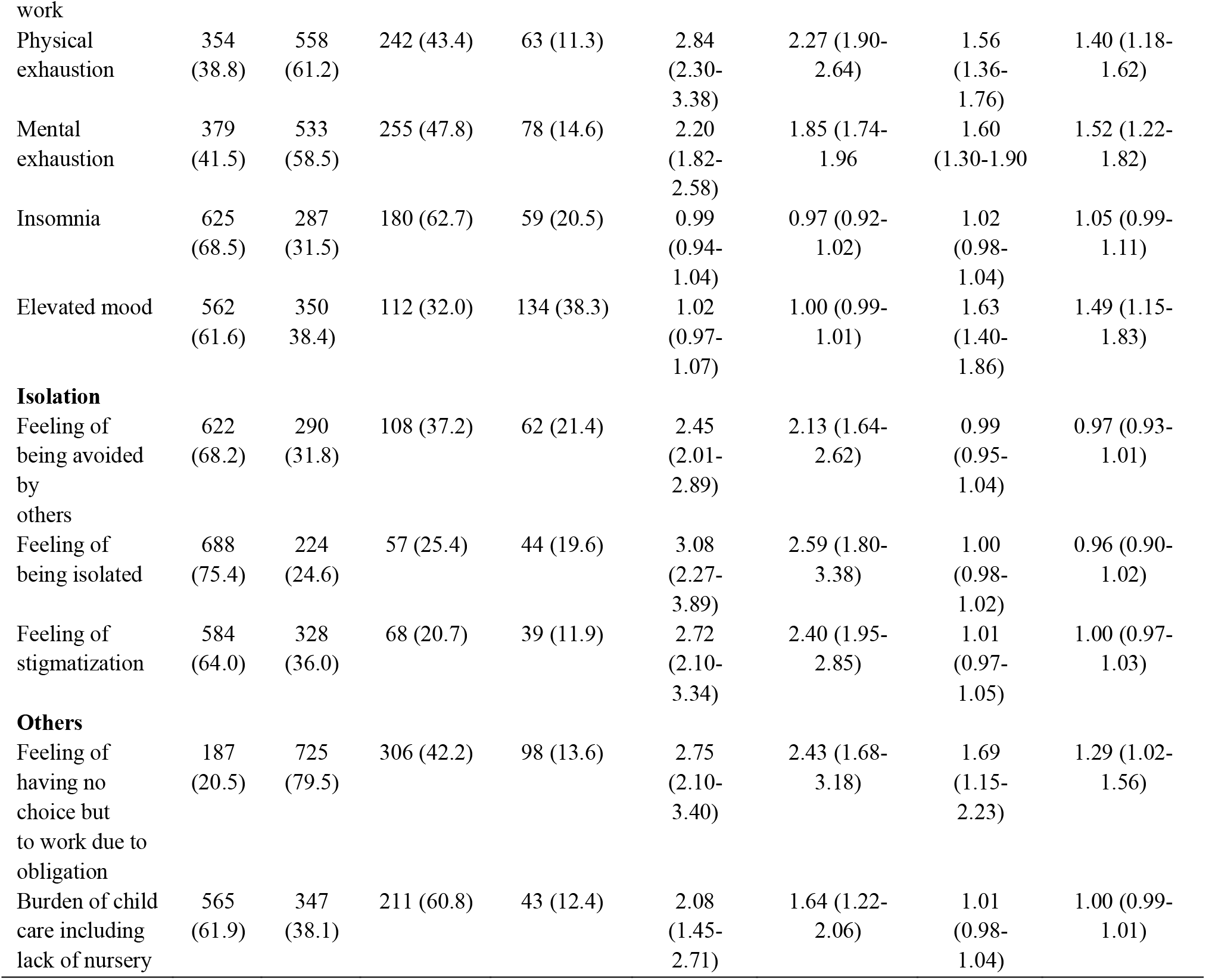
Associations of stress factors and likelihood of reporting hesitation and motivation to work

## Discussion

The present research has explored health workers’ professionalism and willingness to work in a high-risk setting, such as the COVID-19 pandemic in Poland. Thereby, we examined what influences this willingness from the two aspects of motivation and hesitation, revealing that there is a complex mechanism at play. Specifically, there are factors that increase motivation yet do not reduce hesitation, while other factors indeed increase both motivation and hesitation. This study defined four categories of factors based on to what extent they affected motivation and hesitation of healthcare workers: those raising motivation while reducing hesitation; those raising only motivation; those raising only hesitation; and those raising both motivation and hesitation.

This is noteworthy as supporting health workers’ professionalism in a high-risk setting relies on developing a comprehensive awareness of the factors affecting the likelihood of conflict.

Crucial factors to consider are those that mitigate emotional conflict and encourage greater willingness through increased motivation and decreased hesitation. In particular, more effort is required from local and national governments to protect health workers, as well as from hospitals, especially in terms of taking precautions to prevent infection and handling malpractice suits. Although the hospital workers assigned to COVID-19 wards and outpatient areas were given appropriate personal protective equipment, including facemasks, protective suits, and googles, as well as antiviral agents, many felt inadequately protected by their local and national governments and hospitals [18]. In particular, the respondents expressed concerns about a lack of planning on what to do in the event they contracted the disease, as well as a lack of moral support for hospitals from local and national governments. As protection from local and national governments and hospitals raised motivation while reducing hesitation, any positive action targeting these areas is likely to be the most effective in addressing non-illness absenteeism among health workers; thus, hospitals and government should be called upon to consider the importance of such interventions [19, 20]. According to Samuel et al. [21], two factors play a role in inculcating in health workers a sense of duty towards patients during health emergencies. The first relates to the social norm of reciprocity, referring to society’s efforts to take reasonable steps as part of epidemic preparedness to protect health workers and their family members from infection, care for those that do get infected, adequately compensate the families of workers who die after being infected at work, and mitigate malpractice suits for workers in high-risk settings. The second is that health workers’ responsibilities should be lessened, yet not removed, in the effort to protect them from infection, e.g. through a reduction in working hours, reduced caseloads, or reassignment to facilities with less burden or risk. Ensuring that health workers feel safe will enable them to perform their duties, and this effect is strongest when coming from their organizations. However, there must also be consistent communication between both organizations and governments and the health workers, for example in the form of moral support to boost workers’ mental health.

This study found that male and middle-aged health workers showed more motivation and reduced hesitation; this is not in line with previous research in the US (hypothetical influenza pandemic) [22] and Singapore (hypothetical SARS pandemic) [23], which both found no such correlations. This may stem from the fact that middle-aged males held the majority of management positions in our study’s sample population, which tends to imply a strong sense of duty. However, it may be also due to the difference in the contexts: While the prior research focused on hypothetical pandemics, our occurred in the midst of a real one. Meanwhile, previous research has shown that female health workers tend to experience higher levels of stress than males; therefore, additional measures, especially offering childcare services, would help relieve the pressure on female workers [24]. Finally, factors found to raise only motivation may not always have long-term positive benefits as they can eventually lead to burn-out. An incongruous result of this study was that performing unfamiliar work in a high-risk and high-demand working environment leads to enhanced motivation; however, these respondents tended to have less direct patient contact and therefore reduced perceived risk.

It should be noted that only by decreasing the barriers to working in high-risk contexts can a reduction in those stress factors causing hesitation be achieved. These include not knowing about prevention or protection from the disease, the perception that others are avoiding them, and the burdens of increased workload and the lack of available childcare. Some of the measures available to mitigate these factors are work rotation and the sharing of duties, which would not only relieve the workload for individual workers but would also allow health workers to develop more practical knowledge of protection and prevention while alleviating their isolation by allowing them to feel part of the bigger effort.

There should also be a focus on addressing the factors that raise both motivation and hesitation because, even though it may not initially be obvious, the resulting conflict can have a long-lasting impact on health workers. The majority of respondents in this study reported a substantial fear of infection and of passing the infection on to their families, and feeling forced to work. Against the backdrop of a devastating pandemic, it is to some extent natural for those involved to experience feelings of fatigue, mental and physical exhaustion. Nevertheless, research has shown that the most strategies to mitigate these effects are those that incorporate the capacity for health workers and their families to gain preferential access to protective equipment and/or antiviral therapies [25]. While the sample population in this study received these protective measures from their hospitals, these were not extended to their families. However, ensuring their families are similarly protected would go some way towards raising health workers’ motivation and reducing their hesitation. Furthermore, local and national governments and the hospital management should implement strategies to help health workers fulfil their duties as well as ensure they are cared for and compensated should they contract the virus. Doing so would strengthen the perception among health workers that they are protected by hospitals and local and national governments. Finally, it should be noted that although this study was conducted in the context of the COVID-19 pandemic, our findings can to some extent be generalized to other high-risk contexts. Further research is recommended to assess current results’ external validity.

## Limitations and conclusions

A number of limitations of this study should be mentioned. First, the cross-sectional nature of this investigation precludes us from drawing causal inferences. Second, although Internet-based data collection can provide substantial statistical power, the sample of participants may be biased toward those individuals who have access to and regularly use the Internet. Third, we relied exclusively on self-report measures. Keeping these limitations in mind, this study has revealed some of the factors informing health workers’ motivation and hesitation as part of their willingness to work during the COVID-19 pandemic in Poland. Some of these factors increased motivation and hesitation together, thereby having a conflicting effect. Based on these findings, local and national governments as well as hospitals should focus on instilling a sense among health workers that they are sufficiently protected while carrying out their duties, thereby enhancing motivation and reducing hesitation. This can be done through the two-fold approach of ensuring they have access to both protective equipment and compensation while strengthening communication to provide moral support. With the possibility of potentially worse and more lasting pandemics on the horizon, this must be given the highest priority.

## Data Availability

Data are available upon request to the corresponding author.

## Author Contributions

Conceptualization: MM; methodology: MM; statistical analysis: MM; investigation: MM; writing—original draft preparation: MM; writing—review and editing: MM.

## Institutional Review Board Statement

The project was approved by the local ethics committee of the University of Economics and Social Sciences in Warsaw.

## Data Availability Statement

Data are available upon request to the corresponding author.

